# *De novo HDAC3* variants leading to epigenetic machinery dysfunction are associated with a neurodevelopmental disorder

**DOI:** 10.1101/2024.01.29.24301801

**Authors:** Jihoon G. Yoon, Seong-Kyun Lim, Hoseok Seo, Seungbok Lee, Jaeso Cho, Soo Yeon Kim, Hyun Yong Koh, Annapurna H. Poduri, Deciphering Developmental Disorders Study, Jung Min Ko, Dohyun Han, Jong-Hee Chae, Chul-Hwan Lee

## Abstract

Histone deacetylase 3 (HDAC3) is a crucial epigenetic modulator essential for brain development. Although its dysfunction is increasingly recognized in various neurodevelopmental disorders, there have been no reports of human diseases related to HDAC3 dysfunction in Online Mendelian Inheritance in Man (OMIM). This study establishes a novel link between heterozygous *de novo* variants in *HDAC3* and a distinct neurodevelopmental syndrome, characterized by intellectual disability, developmental delays, and other variable manifestations such as musculoskeletal anomalies and congenital heart defects. In a cohort of six individuals, we identified *de novo* missense *HDAC3* variants (D93N, A110T, P201S, L266S, G267S, and R359C), all located in evolutionarily conserved sites. Using trio exome sequencing and extensive phenotypic analysis, we correlated these genetic alterations with the observed clinical spectrum. Our investigations using HDAC assays and western blot analyses identified reduced deacetylation activity in the L266S and G267S variants, positioned near the enzymatic pocket. Additionally, proteomic analysis employing co-immunoprecipitation revealed that disrupted interactions with key multi-protein complexes, particularly CoREST and NCoR in the A110T variant, suggesting a critical pathogenic mechanism. Moreover, immunofluorescence analysis revealed diminished fluorescence intensity (nuclear to cytoplasmic ratio) in the A110T, G267S, and R359C variants, indicating impaired nuclear localization. This study highlights that *de novo HDAC3* variants are associated with a novel neurodevelopmental syndrome, emphasizing the importance of histone deacetylase activity, multi-protein complex interactions, and nuclear localization for normal cellular function of HDAC3. These insights open new possibilities for understanding the molecular mechanisms of this uncharacterized neurodevelopmental disorder and may inform future therapeutic approaches.

**Graphical abstract:** 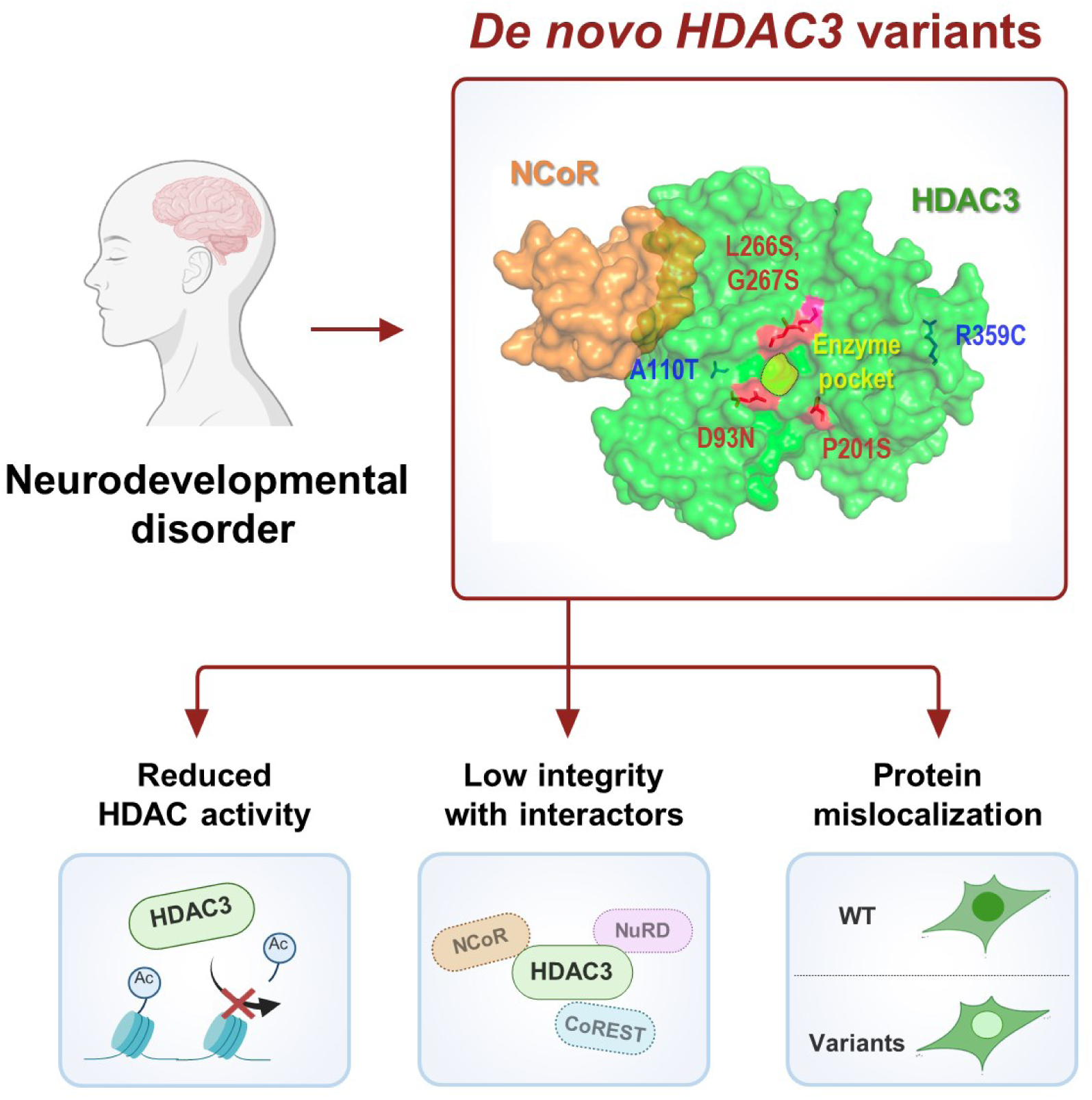

This study identifies *de novo HDAC3* variants in patients with neurodevelopmental disorders, characterized by intellectual disability, developmental delays, and other variable manifestations. We demonstrate that these variants result in reduced HDAC activity, compromised interactions with multi-protein complexes, and improper nuclear localization of the HDAC3 protein. This provides a novel gene-disease association and offers insights into the molecular underpinnings of this disorder.

## Introduction

Epigenetic control of gene expression plays a pivotal role in numerous developmental processes. Among the major epigenetic modifications, histone acetylation is critical, regulated by two classes of proteins: histone acetyltransferases (HATs) and histone deacetylases (HDACs). HDACs remove acetyl groups from lysine (K) residues on histones (H), thereby inducing a closed chromatin structure and transcriptional repression. The deacetylation of histone H3K9 and H3K27 is particularly important, as these lysine residues can be methylated, facilitating chromatin compaction and gene silencing.^1–3^

There are 18 mammalian HDACs classified into four classes (I, II, III, and IV).^4^ HDAC1, HDAC2, HDAC3, and HDAC8 are class I HDACs, primarily nuclear enzymes with strong histone deacetylase activity. These HDACs are often found in multi-protein complexes including the nuclear receptor co-repressor (NCoR) complex, co-repressor of repressor element 1 silencing transcription factor (CoREST) complex, and nucleosome remodeling and deacetylase (NuRD) complex. Notably, the presence of either NCoR1 or NCoR2 (also known as SMRT; silencing mediator of retinoic acid and thyroid hormone receptor) is essential for histone deacetylase activity.^5–7^ HDACs also interact with the CoREST complex, which links to the lysine-specific histone demethylase 1 (KDM1A; also known as LSD1), enzymes responsible for demethylating H3K4 and potentially H3K9, through REST corepressor 1/2 (RCOR1/2).^8–10^ Moreover, HDACs work cooperatively with chromatin remodelers such as CHD3, CHD4, and CHD5 by forming the NuRD complex.^11^

HDAC3, widely present in the cell nucleus and cytoplasm, is recognized as crucial regulators of numerous developmental processes. Studies using HDAC3 mouse models have revealed profound impacts on metabolism and developmental functions in various organs, including the cerebral cortex and cerebellum,^12–14^ heart,^15,16^ and skeletal bone and muscles.^17,18^ It is not only the deacetylation activity that is critical but also the interaction with multi-protein complexes that is vital for brain functions. For instance, NCoR complexes, comprising NCoR1 or NCoR2, are essential in regulating GABA signaling, thereby influencing memory and learning.^19^ Individuals with *de novo* variants in *KDM1A*, which forms an interaction with HDACs via the CoREST complex, present with a neurodevelopmental syndrome characterized by features such as cleft palate, psychomotor retardation, and distinctive facial features (Online Mendelian Inheritance in Man [OMIM] #609132).^20^ Furthermore, *HDAC3* has been implicated in Rett syndrome-like phenotypes through its interaction with methyl CpG binding protein 2 (MECP2), which regulates a subset of neuronal genes through the HDAC3-FOXO deacetylation axis.^14^

To date, several genes in the epigenetic machinery have been associated with Mendelian disorders. Among HDACs, *HDAC4* and *HDAC8* have been established as contributors to Neurodevelopmental disorder with central hypotonia and dysmorphic facies (OMIM #619797) and Cornelia de Lange syndrome 5 (OMIM #300882), respectively.^21,22^ While previous mouse models have robustly supported the association between abnormal brain development and *HDAC3* defects, and one human case has been reported,^19^ further evidence is needed to establish robust gene-disease association and genotype-phenotype relationships in human. In this study, we described the phenotypes of individuals with *de novo* heterozygous variants in *HDAC3* and characterized the functional impacts of the identified *de novo* variants.

## Materials and methods

### Subjects

Patients were enrolled from the Rare Disease Center of Seoul National University Hospital (SNUH), Seoul, Republic of Korea, the Boston Children’s Hospital (BCH) Rare Disease Cohorts Initiative,^23^ or Deciphering Developmental Disorders (DDD) study.^24^ Appropriate informed consent was obtained for patients, including for the publication of all photographic material as appropriate. The study received approval from the Institutional Review Boards of the participating institutions in compliance with the Declaration of Helsinki (SNUH IRB No. 1406-081-588).

### Sequencing analysis

Blood samples were collected from trios (proband and parents), and genomic DNA was subjected to Illumina short-read sequencing. Detailed methods for exome sequencing have been previously documented.^25^ Sequenced reads were aligned to the human reference genome hg38 and processed using the Exome Germline Single Sample pipeline (v3.0.4), as provided by the WDL Analysis Research Pipelines (WARP; Broad Institute, MA, USA). Detected *de novo* variants in *HDAC3* were subsequently validated through Sanger sequencing (**Figure S1**) with PCR experiments using the target-specific primers (**Table S1**).

### Protein modeling

The protein structure of the NCoR2 DAD domain-HDAC3 complex was retrieved from the Protein Data Bank (PDB: 4A69).^26^ Subsequent structural adjustment and three-dimensional visualization were performed using PyMOL (v2.5.5; PyMOL Molecular Graphics System, Schrödinger Inc., New York, NY, USA).

### Mutagenesis of HDAC3 variants

To generate *HDAC3* variants, we performed PCR with mutagenic primers (Macrogen, Seoul, Korea; **Table S1**) using nPfu-Forte polymerase (Enzynomics, Daejeon, Korea). After PCR reaction, PCR products were digested with DpnI restriction enzyme (Enzynomics, Daejeon, Korea) at 37℃ for 1hr. HDAC3 variant clones were transformed into DH5α competent cells and confirmed by Sanger sequencing (**Figure S2B**).

### Purification of proteins using baculovirus expression system

To purify the complexes of the human NCoR1 DAD domain and HDAC3 (**Figure S2C**), 6ⅹHis-tagged NCoR1 DAD domain and FLAG-tagged HDAC3 were cloned into the baculovirus expression vector, pFastBac1 (Invitrogen, Cat# 10-360-014). The NCoR1 DAD domain, HDAC3 wild-type (WT), or variants were co-expressed in Sf9 cells (Life Technologies, Cat# 11496-015) through baculovirus infection. After 64hrs of infection, Sf9 cells were harvested in BC350 buffer (20 mM HEPES-NaCl at pH 7.8, 350 mM NaCl, 10% Glycerol, 0.1% NP-40, 1 mM EDTA) with protease inhibitors (1 mM Phenylmethylsulfonyl fluoride [PMSF], 0.1 mM Benzamidine hydrochloride, 1.25 mg/ml Leupeptin, 0.625 mg/ml Pepstatin A) and phosphatase inhibitors (20 mM NaF, 1 mM Na_3_VO_4_). Cells were lysed by sonication, and the WT or variant recombinants were purified by Ni-NTA Agarose (QIAGEN) and Anti-FLAG M2 Affinity Gel (Sigma). The NCoR1 DAD domain (Clone ID: KU017294) and HDAC3 plasmids (Clone ID: hMU006316) were provided from the Korea Human Gene Bank (Medical Genomics Research Center, KRIBB, Korea).

### HDAC assay

HDAC assays were performed with HDAC buffer (10 mM Tris-HCl at pH 8.0, 150 mM NaCl, 10% Glycerol), recombinant H3K27-acetylated mononucleosomes or recombinant H3K4,9,14,18/H4K5,8,12,16-acetylated mononucleosomes (EpiCypher), and recombinant human NCoR1 DAD domain-HDAC3 complexes. The reaction proceeded for 60 min at 30℃ and stopped with the addition of 4μl of sample buffer (50 mM Tris-HCl at pH 6.8, 2% SDS, 6% Glycerol, 0.004% Bromophenol blue, 5% β-mercaptoethanol). HDAC activities were measured with H3K27ac, H3ac, and H4ac antibodies (**Table S2**, **Figure S3**).

### Cell culture, lentiviral production and transduction, and lysate preparation

Human embryonic kidney (HEK) 293T cells (Cat# CRL-3216) were obtained from the American Type Culture Collection (ATCC, USA). HEK293T cells were maintained in Dulbecco’s Modified Eagle’s Medium (DMEM; Biowest) supplemented with 5% (v/v) fetal bovine serum (FBS) and 1x Antibiotic-Antimycotic (Gibco) in a humidified atmosphere of 5% CO_2_ at 37°C. HDAC3 WT or variant plasmids were subcloned into the pLV-EF1α-IRES-Puro vector (Addgene, #85132) for lentiviral production. Lentiviral vectors containing HDAC3 WT or variants at a concentration of 10μg each were co-transfected with 2.5μg of pcREV, 3μg of BH10, 5μg of VSVG packaging vector into HEK293T cells. The viral supernatant was harvested 48hrs after transfection and used to infect target cells, which were plated in 6-well plates. Infected cells were selected with 2μg/ml puromycin for five days, and the medium containing puromycin was refreshed daily. To prepare cell lysates, 8.8ⅹ10^6^ cells were washed with ice-cold PBS and subsequently lysed using RIPA buffer (50 mM Tris-HCl at pH 8.0, 400 mM NaCl, 1% NP-40, 0.5% sodium deoxycholate, 0.1% SDS) along with protease inhibitors, as well as phosphatase inhibitors. The cells were subjected to sonication for lysis and subsequently clarified by centrifugation at 13,000 rpm, 4℃ for 20 min. The total protein concentration was measured by Pierce BCA Assay Kit (Thermo Fisher Scientific).

### Western blotting

Cell lysates were prepared by quantifying and normalizing protein concentrations, followed by denaturation of 10-15µg of total protein in sample buffer (50 mM Tris-HCl, pH 6.8, 2% SDS, 6% glycerol, 0.004% bromophenol blue, 5% β-mercaptoethanol) at 95°C for 5 min. The samples were then resolved by SDS-PAGE, running initially at 100V for 15 min and subsequently at 120V for 75 min. Proteins were transferred onto PVDF membranes (Merck Millipore), which were subsequently blocked with 5% skim milk in TBS containing 0.1% Tween 20 (TBS-T) at RT for 1hr. The membranes were incubated overnight at 4°C with the primary antibodies. Following primary incubation, membranes were rinsed three times for 10 min each with TBS-T at RT. Next, the membranes were probed with HRP-conjugated secondary antibodies at RT for 1hr. After secondary antibody incubation, the membranes underwent three additional 5-minute washes with TBS-T. Detection was performed using an enhanced chemiluminescent substrate (Thermo Fisher Scientific). The antibodies utilized are specified in **Table S2**.

### Co-immunoprecipitation

A total of 1.5mg of cell lysate was prepared with BC150 buffer composed of 20 mM HEPES-NaCl, pH 7.8, 150 mM NaCl, 10% glycerol, 0.1% NP-40, and 1 mM EDTA, supplemented with protease inhibitors and phosphatase inhibitors. The lysate was then incubated with 50 µl of Anti-FLAG M2 Affinity Gel (Sigma) at 4°C for 3hrs. Subsequent to incubation, the gel was washed three times with BC150 buffer at 4°C for 10 min per wash. Following the final wash, the complexes were eluted using 0.2mg/ml 3ⅹFLAG peptide at 4°C for 1hr. The eluted proteins were resolved on SDS-PAGE gels, which were subsequently either silver-stained or electro-transferred onto 0.45 µm PVDF membranes (Merck Millipore) for western blot analysis.

### Silver staining

The SDS-PAGE gel was fixed using a fixative solution containing methanol. The gel was immersed in this solution at RT for 20 min, the procedure repeated twice, and subsequently rinsed twice with distilled water for 10 min each. The gel was then stained with a solution containing NaOH, ammonium hydroxide, and silver nitrate (CHEMICALS DUKSAN, Incheon, Korea) at RT for 15 min. Following the staining, it was rinsed twice with distilled water and developed in a solution of citric acid and formaldehyde at RT for a duration ranging from 2 to 20 min, depending on the desired intensity of staining.

### Mass spectrometry (MS) analysis

Proteomic analysis was performed through an optimized protocol integrating filter-aided sample preparation and StageTip desalting, as previously established.^27^ Protein samples were initially denatured using a buffer containing 2% sodium dodecyl sulfate (SDS), 50 mM chloroacetamide, and 10 mM tris(2-carboxyethyl)phosphine hydrochloride in a 0.1 M Tris-HCl solution (pH 8.5). The mixture underwent reduction and alkylation processes at 95°C for 15 min. Digestion was facilitated overnight at 37°C with a trypsin/Lys-C mix at a 1:100 protease-to-protein mass ratio. Post-digestion, peptides were acidified with 10% trifluoroacetic acid (TFA) and purified using StageTip columns, with styrene-divinylbenzene-reverse phase sulfonate as the adsorbent.

The peptides were then separated and analyzed on a Q-Exactive HF-X mass spectrometer (Thermo Fisher Scientific, Waltham, MA, USA) coupled with an Ultimate 3000 RSLCnano system (Dionex, Sunnyvale, CA, USA).^28^ A comprehensive dual-column setup, comprising a C18 trapping column and an EASY-Spray C18 analytical column, was utilized for peptide separation. A 120-minute gradient, ranging from 5% to 40% acetonitrile (ACN), was employed at a 300 nl/min flow rate. MS detection was operated in positive ion mode, performing an initial full scan across an m/z range of 300 to 1,800 at 60,000 resolution, followed by MS/MS scans at 15,000 resolution for the top 15 precursor ions, selected within a 1.6 m/z isolation window and fragmented with a normalized collision energy of 30%.

Data processing was conducted using MaxQuant (v2.2.0.0; Max Planck Institute of Biochemistry, Munich, Germany),^29^ with MS/MS spectra matched against the Human UniProtKB/Swiss-Prot database (June 2023 release; Homo sapiens, 20,423 entries) complemented by four HDAC3 variants and common contaminants. A 6ppm tolerance was applied to the precursor ion, with 20ppm for MS/MS ions. Fixed and variable modifications included carbamidomethylation of cysteine and, respectively, N-terminal acetylation and methionine oxidation. A stringent 1% false discovery rate was maintained for all peptide and protein identifications. Label-free quantification leveraged the iBAQ algorithm,^30^ facilitating robust absolute quantification within the MaxQuant framework.

### Immunofluorescence staining

HEK293T cells were seeded onto Cell Culture Slide I (SPL, Gyeonggi-do, Korea). The cells were fixed with 4% paraformaldehyde (v/v), for 30 min and then permeabilized with 0.1% triton X-100 for 20 min. Nonspecific antibody binding sites were blocked by incubation with 1% BSA in PBS, for 20 min. The cells were then incubated overnight at 4℃ with primary antibody specific to FLAG M2, followed by washing with PBS three times. An anti-mouse IgG-Alexa Fluor 488 secondary antibody was added to the cells and incubated for 2hrs at RT. Cell culture slides were then washed, and mounted by applying Antifade Mounting Medium (Vectashield). Fluorescence images were acquired using a confocal laser-scanning microscope and software (FV31S-SW v2.3), with a 60X objective (Olympus FV3000, Tokyo, Japan). DAPI staining was performed for nuclear staining.

### Statistical analysis

Our analysis employed two-sample *t*-tests to compare protein interaction alterations between HDAC3 WT and variants. Significance was assigned to proteins displaying a minimum fold change (FC) of 1.5 coupled with a *P* value less than 0.05. We utilized Perseus software (v2.0.11) for the statistical analysis of the MS data.^31^ To discern potential interaction partners, we processed the iBAQ intensities through a log2 transformation. Proteins were considered for further analysis if they exhibited at least two-thirds valid data points within each experimental condition. We addressed missing values using the deterministic minimal value (MinDet) imputation method,^32^ which aids in mitigating the bias that can arise from non-random data absence. ImageJ (v1.54) was used for the quantification of western blot and analysis of immunofluorescent images. Statistical analysis was performed using GraphPad Prism (v9.5.0).

## Results

### Identified *de novo* variants and associated clinical findings

We first identified two unrelated females with *de novo HDAC3* variants, who were previously undiagnosed with exome negative results and presented with neurodevelopmental disorders characterized by a spectrum of clinical manifestations. These variants, resulting in the A110T and R359C substitutions, were validated through Sanger sequencing as depicted in **Figure S1**. An additional case with the P201S variant, was identified from an epilepsy cohort of 522 cases.^23^ Analysis of 13,451 individuals enrolled in the Deciphering Developmental Disorders (DDD) cohort further revealed three additional heterozygous *de novo* HDAC3 variants (D93N, L266S, and G267S),^24^ as outlined in **Table 1**. All variants were missense variants located at evolutionarily conserved sites in vertebrates (**Figure 1A**). Structural examination with the previously reported HDAC3 and NCoR2 deacetylase activating domain (DAD) domain complex (**Figure 1B**) demonstrated that the D93N, P201S, L266S, and G267S variants were proximal to the enzyme’s active catalytic sites (H134 and H135).^26^ The A110T variant was adjacent to the interface with the NCoR2 DAD domain, whereas the R359C variant was positioned on an alpha-helix away from the catalytic center and interaction domain. Notably, the R359 residue engages with the H234 residue, which may be integral to the stability of the complex, and situated within the domain identified as the nuclear localization signal (NLS), as previously defined through domain mapping **(Figure 1A)**.^33,34^ Additionally, NCoR1 and NCoR2 had high sequence similarities in the HDAC3 binding sites (**Figure S2A**).

**Figure 1.**
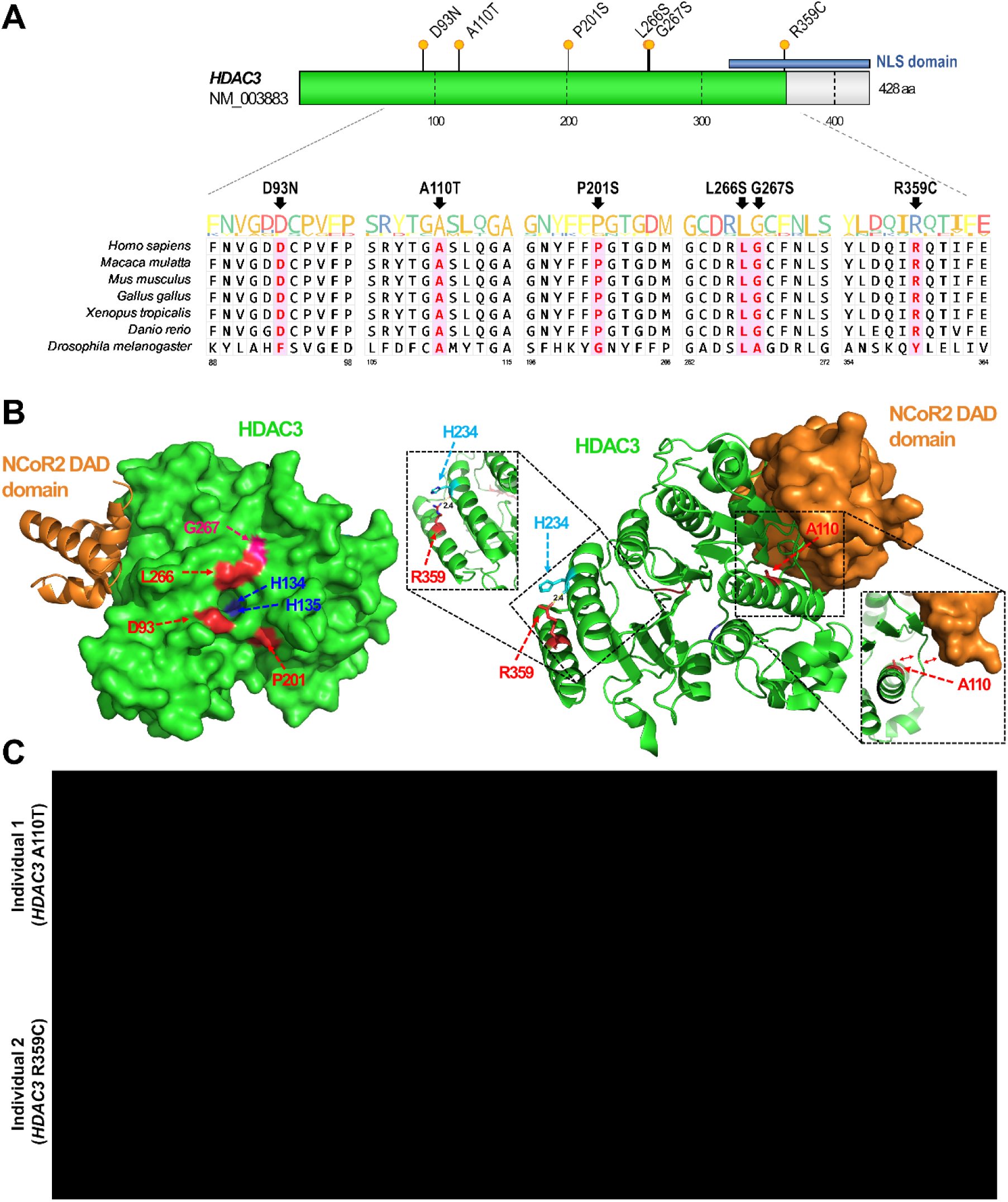
Locus conservation, three-dimensional structure, and phenotypic characteristics of individuals with *de novo HDAC3* variants. **(A)** Genomic locations of identified *de novo* missense variants. All variants are situated in highly conserved regions across vertebrate species. The blue box indicates the presence of the nuclear localization signal (NLS) domain. **(B)** Three-dimensional structure of HDAC3-NCoR2 DAD and variant locations (modified from PDB: 4A69). This figure provides a three-dimensional representation of the HDAC3 protein structure, highlighting the locations of the six identified HDAC3 variants (D93N, A110T, P201S, L266S, G267S, and R359C). **(C)** Phenotypic characteristics of individuals 1 and 2 carrying the A110T and R359C variants, respectively. Both patients exhibited facial dysmorphism, profound microcephaly with no abnormal brain MRI findings, and skeletal abnormalities such as joint stiffness, scoliosis, or polysyndactyly (yellow triangle).

**Table 1.** Genetic and clinical findings in individuals with *de novo HDAC3* variants.

| Clinical findings | Individual 1 | Individual 2 | Individual 3 | Individual 4 | Individual 5 | Individual 6 |
| --- | --- | --- | --- | --- | --- | --- |
| **Sex** | F | F | M | F | M | NA |
| **Age at evaluation** | 21–25 | 0–5 | 11–15 | 11–15 | NA | NA |
| **HDAC3 *de novo* variant (NM_003883.4)** | c.328G>A, p.(Ala110Thr) | c.1075C>T, p.(Arg359Cys) | c.601C>T, p.(Pro201Ser) | c.797T>C, p.(Leu266Ser) | c.277G>A, p.(Asp93Asn) | c.799G>A, p.(Gly267Ser) |
| **CADD score (Phred)** | likely deleterious; 27.3 | probably deleterious; 32 | likely deleterious; 28.3 | likely deleterious; 29.9 | likely deleterious; 27.3 | likely deleterious; 27.5 |
| **Conservation score (GERP++_RS)** | 4.45 | 5.38 | 5.45 | 5.37 | 5.38 | 5.37 |
| **Facial dysmorphism** | Yes | Yes | No | NA | NA | NA |
| **Developmental delay** | Yes | Yes | No | Yes | Yes | Yes |
| **Failure to thrive** | Yes | Yes | No | NA | NA | NA |
| **Epilepsy** | Yes | No | Single seizure, abnormal EEG | NA | NA | NA |
| **Intellectual disability** | Yes | Yes | No | Yes | Yes | Yes |
| **Autistic behavior** | Yes | No | No | NA | NA | NA |
| **Brain MRI findings** | No specific findings | No specific findings | No specific findings | focus of nodular heterotopia | NA | NA |
| **Musculoskeletal abnormalities** | scoliosis, varus of the proximal tibia, hand joint deformity | polydactyly, syndactyly | No | joint hypermobility | No | Yes |
| **Congenital heart defect** | atrial septal defect | atrial septal defect, patent ductus arteriosus | No | No | No | Yes |
| **Others** | sensorineural hearing loss, microptia | congenital hydronephrosis | neonatal torticollis | subcutaneous scalp arteriovenous malformation | NA | Abnormality of ear and eye |
| **Reference** | This report | This report | Koh et al.<sup>23</sup> | Zhou et al.<sup>19</sup>, Kaplanis et al.<sup>24</sup> | Kaplanis et al.<sup>24</sup> | Kaplanis et al.<sup>24</sup> |

Abbreviation: CADD, Combined Annotation Dependent Depletion; GERP, Genomic Evolutionary Rate Profiling; NA, not available;

The clinical profile of the patients is summarized in **Table 1** (detailed descriptions of clinical information have been omitted in accordance with medRxiv guidelines). All patients commonly present with neurodevelopmental problems, including developmental delay and intellectual disability.

### Changes in the histone deacetylase activities

To investigate the impact of HDAC3 variants on histone deacetylase activity, we ectopically expressed HDAC3 WT or four variants (A110T, G267S, L266S, or R359C; **Figure S2B**) in HEK293T cells **(Figure 2A)**. Note that the selection of G267S and L266S variants from G267S, L267S, D93N, and P201S was based on their representation of the variants located in close proximity to the catalytic site **(Figure 1B)**. The G267S and L266S variants, which are located near the catalytic pocket, were found to significantly increase the overall levels of histone acetylation at H3K9ac, H3K27ac, and H3ac **(Figure 2A,B)**. These findings suggest that the G267S and L266S variants substantially impair HDAC activity. In contrast, the A110T and R359C variants did not exhibit noticeable deficiencies in overall HDAC activities.

**Figure 2.**
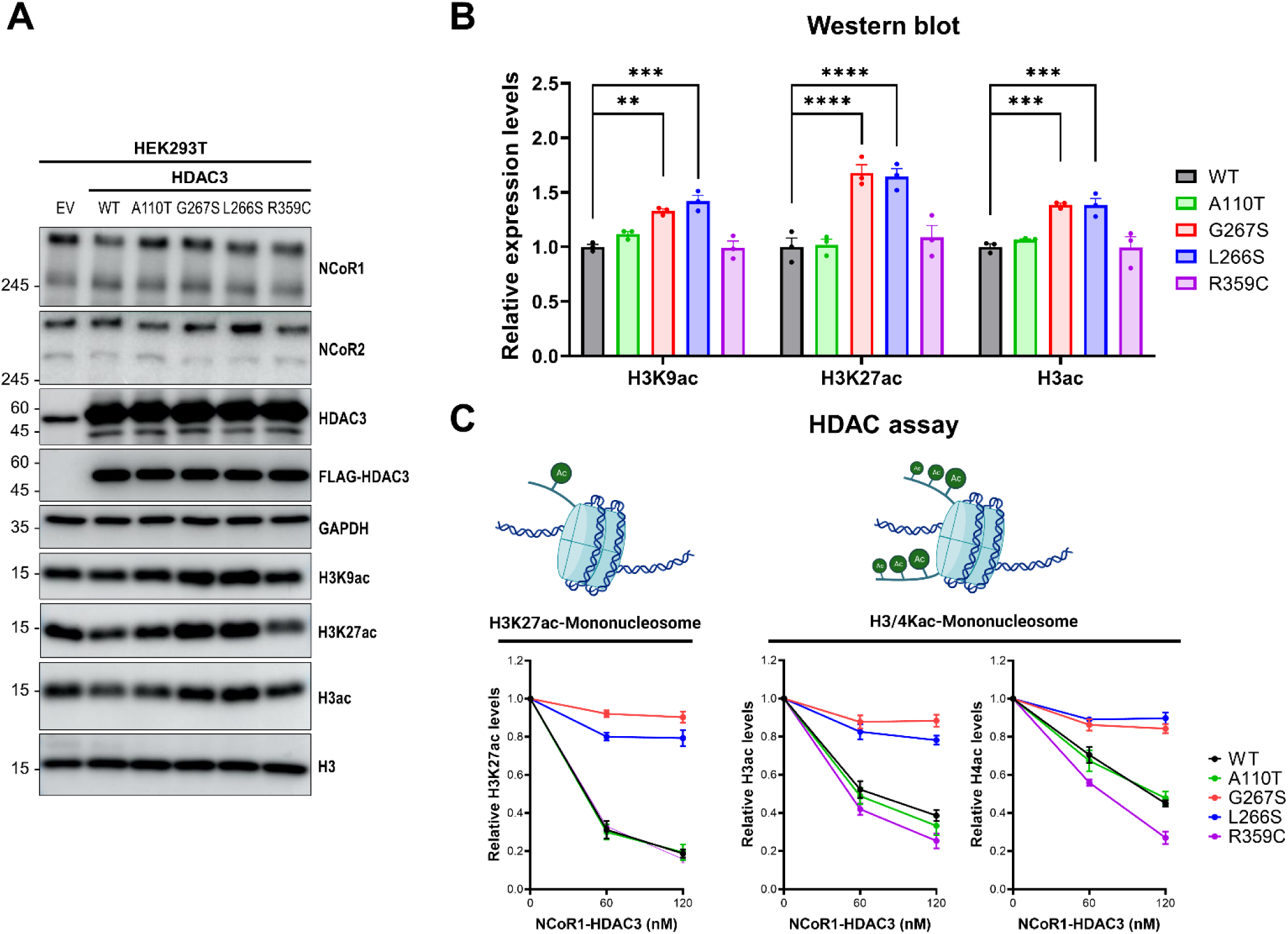
HDAC3 variants near catalytic sites display deficient histone deacetylase activity. **(A)** Western blot analyses in HEK293T cells overexpressing empty vector (EV), FLAG-tagged wild-type HDAC3 (WT), or selected HDAC3 variants (A110T, G267S, L266S, or R359C). Detection of NCOR1, NCOR2, HDAC3, FLAG-HDAC3, GAPDH (loading control), and acetylated histones of H3K9, H3K27, total H3, and H4. **(B)** Quantification of acetylation at histone sites H3K9, H3K27, H3, and H4 from Panel A. The G267S and L266S variants show significantly increased acetylation at H3K9, H3K27, H3, and H4, indicating impaired deacetylation function. \**P* < 0.05, \*\*\**P* < 0.001, \*\*\*\**P* < 0.0001. **(C)** HDAC assays measuring deacetylation of histone peptides H3K27ac, H3ac, and H4ac by complexes of either NCoR1 DAD domain-HDAC3 WT or variant forms. The assays were conducted at various concentrations (0, 60, 120 nM) along with 100nM of acetylated mononucleosomes. The G267S and L266S variants demonstrate notably reduced HDAC activity, contrasting with other variants which align with WT activity.

To further validate whether the HDAC3 variants directly influence histone deacetylase activity, we purified WT and variant HDAC3 proteins in combination with the NCoR DAD domain, which is the minimal requirement for optimal HDAC activity, using baculovirus expression system.^6,7^ The HDAC3 variants did not exhibit any defects in their interaction with the NCoR1 DAD domain (**Figure S2C**). Notably, the interaction with the full-length NCoR1 may have defect in the cell (see below). Subsequently, we performed HDAC assays in triplicate using either mono-nucleosomes acetylated at H3K27 or mono-nucleosomes with acetylations on H3 and H4 **(Figure 2C, Figure S3)**. Consistent with the findings from the *ex vivo* experiments, the combination of NCoR1 DAD domain with either HDAC3-G267S or -L266S variant, displayed a notable impairment on HDAC activity. Similarly, when NCoR1 DAD domain was paired with either HDAC3-A110T or -R359C, no deficiencies in HDAC activity were observed.

### Compromised protein interactions

To further investigate the underlying mechanism for the HDAC3 variants with retained HDAC activity, we explored potential changes in protein-protein interactions. Proteomic analysis was conducted on co-immunoprecipitated samples from HEK293T cells expressing HDAC3 WT, and the four variant constructs. We normalized protein abundance to WT levels, using fold change (FC) and intensity based absolute quantification (iBAQ) as a quantitative measurement. Volcano plot analysis (**Figure 3A**) revealed a downregulation of proteins in the variant forms such as NCoR1/2, TBL1X/Y, KDM1A, RCOR1/3, GSE1, and HDAC1, which are associated with the NCoR, CoREST, and NuRD complexes, while proteins like PSMA7/8, PSMC1, and PSMD14, involved in proteasome degradation pathways, were upregulated (**Figure 3B, 3C**). A universal disruption in the association between RCOR and KDM1A was observed across all HDAC3 variants, implicating a pivotal role for this interaction in maintaining CoREST complex integrity. Furthermore, attenuation of the NCoR complex association was most pronounced with the A110T variant, while the L266S variant demonstrated a compromised affinity for NuRD complex constituents, specifically HDAC1 and RBBP4/7. In contrast, upregulation of the proteasome pathways suggested that the variant forms may be more susceptible to proteasome-mediated degradation due to their impaired interactions with key multi-protein complexes. Particulary, quantified protein levels in iBAQ intensity showed remarkable decrease in NCoR1/2 and KDM1A (**Figure 3D**), which are core molecules of the NCoR and CoREST complexes.

**Figure 3.**
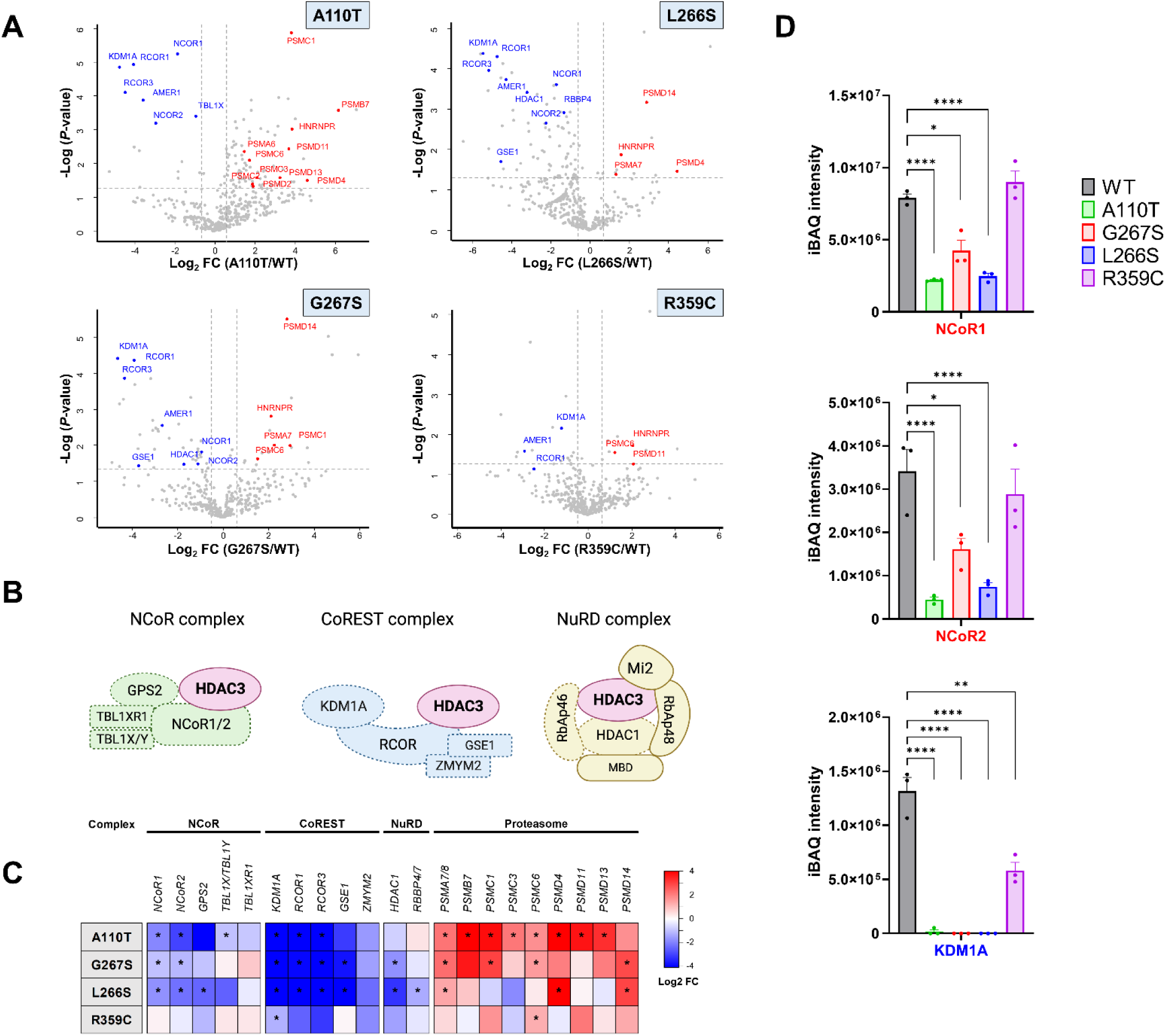
Protein-interactome analysis reveals reduced associations between HDAC3 variants and the subunits of NCoR, CoREST, and NuRD complexes. **(A)** Volcano plots contrasting protein interaction profiles of HDAC3 variants (A110T, G267S, L266S, R359C) to wild-type (WT) HDAC3 in HEK293T cells. The plots highlight proteins with significantly altered interactions, with the x-axis representing the Log2 fold change (FC) and the y-axis showing the -Log10 *P*-value. **(B)** Schematic diagrams of the NCoR, CoREST, and NuRD complexes, with the position of HDAC3 and its interacting subunits within each complex. These diagrams provide a visual summary of the multi-protein complexes relevant to the study. **(C)** A heatmap summarizes the log2 FC in protein interactions for HDAC3 variants relative to WT. Each column represents a distinct protein, as highlighted in Panel A, and each row corresponds to the various HDAC3 variants. Shades of blue indicate a decrease, and shades of red an increase in interaction strength, with asterisks denoting statistically significant changes (*P* < 0.05). The data underscore decreased interactions with subunits of the NCoR, CoREST, and NuRD complexes and contrasted with an increased interaction tendency with subunits of the proteasome pathway, suggesting variant-specific effects on HDAC3 function and complex integrity. **(D)** Bar graphs presenting the intensity based absolute quantification (iBAQ) for selected proteins: NCOR1, NCOR2, and KDM1A. The data reveal a consistent reduction in association with the HDAC3 variants. \**P* < 0.05, \*\**P* < 0.01, \*\*\*\**P* < 0.0001.

To corroborate these proteomic findings, we employed silver staining and western blot analyses. Immunoprecipitation assays in HEK293T cells expressing either the WT or variant forms of HDAC3 delineated distinct protein interaction profiles visualized by silver staining (**Figure 4A**). The A110T variant, in particular, displayed a diminished band correlating with NCoR complex interaction, and a universally reduced interaction with KDM1A was evident across all variants. Western blot analysis further substantiated the significant disruption of the A110T variant’s interactions with full-length NCoR1 and NCoR2 (**Figure 4B,C**). Additionally, the L266S variant presented more severe impairments than the G267S variant with these proteins, consistent with our proteomic analysis (**Figure 3C**). Furthermore, all HDAC3 variants showed diminished interactions with KDM1A. Collectively, our findings demonstrate that HDAC3 variants perturb interactions with CoREST, NCoR, and NuRD complexes. Of particular note is the disrupted interaction with the CoREST and NCoR complexes involving KDM1A and NCoR1/2, which we posit as a central pathogenic mechanism underlying the phenotypes observed in affected individuals.

**Figure 4.**
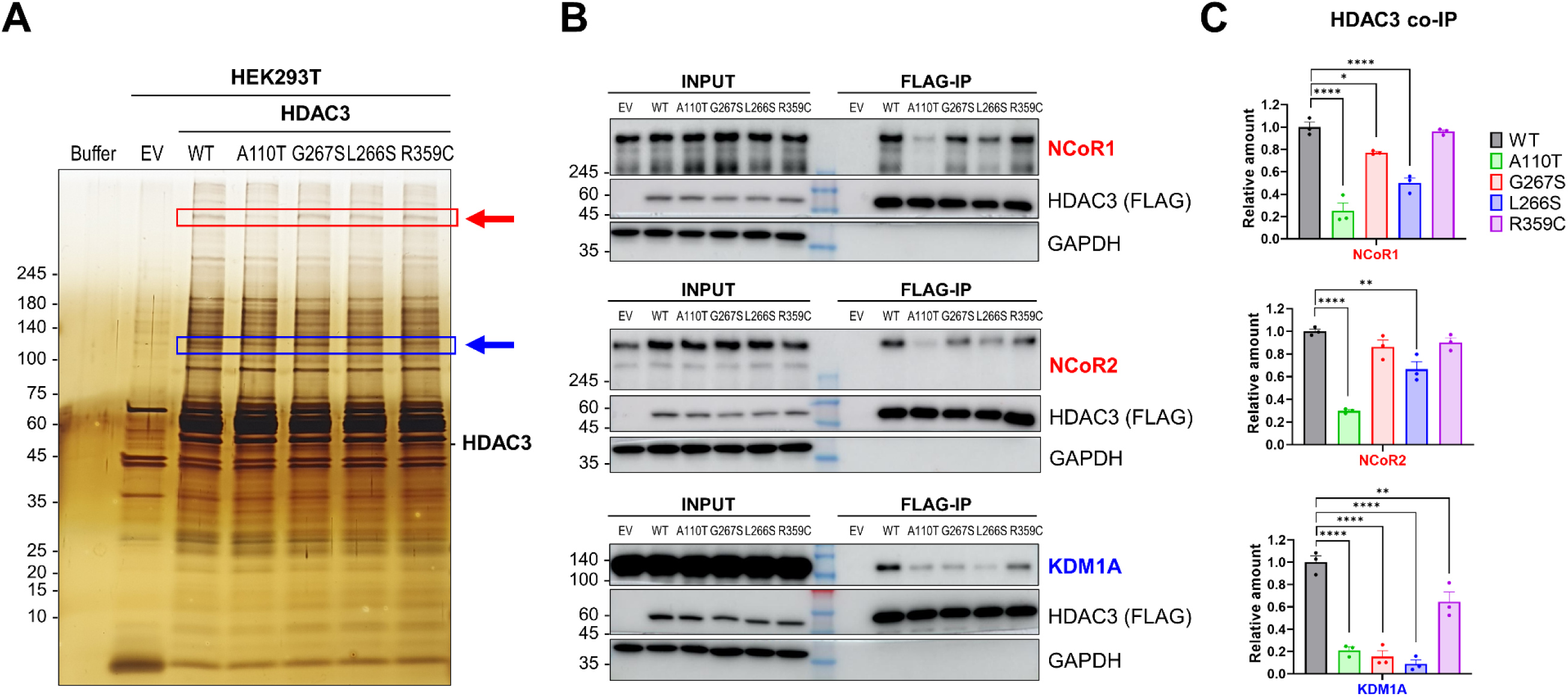
Impaired interactions of HDAC3 variants with NCoR1/2 and KDM1A. **(A)** Silver-stained SDS-PAGE analysis demonstrating the protein complexes co-immunoprecipitated with FLAG-tagged HDAC3 from HEK293T cells. Variants tested include empty vector (EV), wild-type (WT), and A110T, G267S, L266S, R359C HDAC3 variants. The intensity of the bands corresponding to the NCoR complex (red arrow) and KDM1A (blue arrow) is reduced in the variants, with the A110T variant showing a marked decrease in NCoR complex band intensity (red rectangle) and all variants exhibiting diminished KDM1A bands (blue rectangle). **(B)** Western blot assays confirming the differential co-immunoprecipitation of NCoR1, NCoR2, and KDM1A with HDAC3 variants, using an anti-FLAG antibody for immunoprecipitation. FLAG-tagged HDAC3 and GAPDH serve as a reference for protein expression and loading control, respectively. **(C)** Quantification of co-immunoprecipitated NCoR1, NCoR2, and KDM1A, normalized to WT HDAC3 levels, as derived from the Western blot data in (B). This panel quantitatively depicts the interaction deficits of the A110T and R359C variants with NCoR1, NCoR2, and KDM1A, which are significant despite these variants retaining deacetylase activity comparable to WT. \*\**P* < 0.01, \*\*\*\**P* < 0.0001.

### Diminished nuclear localization

In our examination of subcellular protein distribution using fluorescence microscopy, we investigated the localization of HDAC3 within HEK293T cells (**Figure 5A**). FLAG-tagged WT and variant HDAC3 proteins were ectopically expressed, and their localization was detected with an anti-FLAG antibody, while DAPI staining delineated the nuclear boundary. HDAC3 was predominantly localized in the nucleus of the WT cells, whereas the variant forms displayed a discernible reduction in nuclear localization. Quantitative analysis of immunofluorescent intensities, represented as the nuclear-to-cytoplasmic (N/C) ratio (median), revealed a significant decrease in nuclear signal for the A110T (0.67), L266S (0.92), and R359C (1.55) variants compared to the WT (2.03) cells **(Figure 5B**). This reduction is consistent with our proteomic data (**Figure 3**), which showed diminished interactions with components of the NCoR complex, potentially leading to decreased nuclear stability and increased proteasome degradation, particularly for the A110T and L266S variants. Moreover, the R359C variant lies within the nuclear localization signal sequence (amino acids 313-428 of HDAC3).^33,34^ The R359C variant could therefore undermine nuclear localization, resulting in a lower N/C ratio compared to WT cells. Furthermore, the G267S variant, with a median N/C ratio of 1.95, exhibits a nuclear localization similar to WT, although the broad distribution within the cell population may indicate small portions of mildly defective cells. These findings not only underscore the potential pathogenicity of HDAC3 variants but also emphasize protein mislocalization as a critical pathogenic mechanism, further elucidating the consequences of impaired protein interactions within cellular compartments.

**Figure 5.**
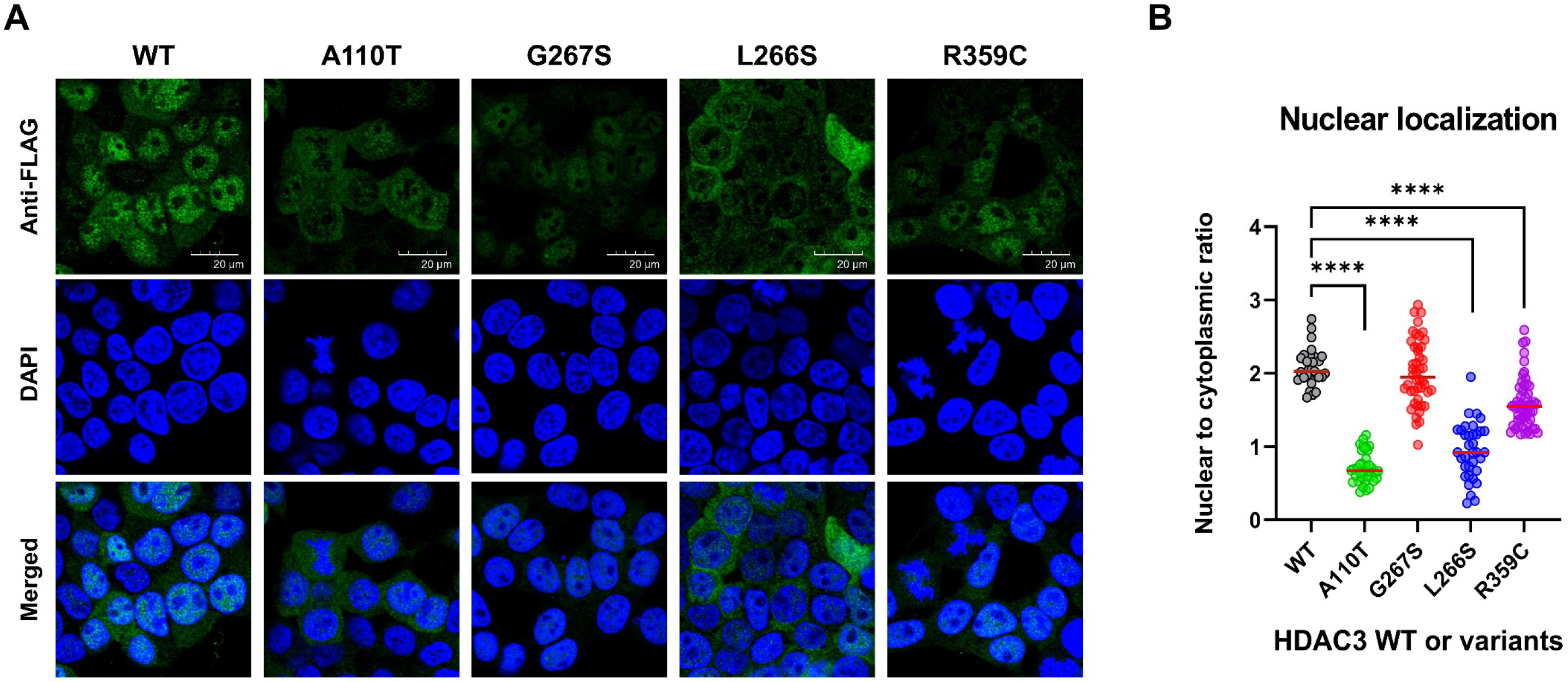
Differential nuclear localization of HDAC3 variants in HEK293T cells. **(A)** Immunofluorescence analysis of HEK293T cells transduced with wild-type (WT) or variant forms of HDAC3 lentivirus. Cells were stained with an anti-FLAG antibody to visualize HDAC3 or its variant forms (green) and with DAPI to mark DNA (blue). HDAC3 WT prominently localizes in the nucleus, as shown by intense green fluorescence. In contrast, the A110T and L266S variants show notably weaker nuclear fluorescence and stronger cytoplasmic fluorescence, implying compromised nuclear localization of HDAC3. The G267S variant exhibits relatively conserved nuclear localization similar to the WT. **(B)** Quantitative assessment of nuclear localization. This panel presents the quantification of nuclear versus cytoplasmic fluorescence intensity, expressed as the nuclear-to-cytoplasmic (N/C) ratio, in cells transfected with HDAC3 WT or variants of HDAC3. Individual data points correspond to measurements from single cells, with the median value of each variant depicted by the horizontal red line. The N/C ratios for A110T, L266S, and R359C variants are significantly reduced in comparison to the HDAC3 WT, indicating a deficiency in nuclear accumulation. Conversely, the G267S variant exhibits a nuclear localization similar to WT, though the broad spread of data points indicates variable localization within the population. Statistical significance is indicated by asterisks, with \*\*\*\**P* < 0.0001 denoting a highly significant difference from the WT ratio.

## Discussion

This study elucidates the association of *HDAC3* with a novel neurodevelopmental syndrome, characterized by a constellation of clinical features including intellectual disability, congenital heart defects, and skeletal abnormalities, as evidenced in six unrelated patients carrying heterozygous *de novo* variants (**Table 1**). Each identified variant was missense, occurring at evolutionarily conserved sites, and predicted to be deleterious through multiple *in silico* tools. We observed that the L266S and G267S variants, located near the active enzyme pocket, have reduced catalytic activities, while A110T and R359C have intact catalytic activities with NCoR DAD domain, as determined by *in vitro* functional assays. Further exploration using co-immunoprecipitation and protein interactome analysis demonstrated a consistent disruption of interaction with *KDM1A*, a key constituent of the CoREST complex, across all variants tested. Moreover, disruption in interactions with other multi-protein complexes, such as the NCoR complex and the NuRD complex, was also observed in specific variants, suggesting that specific variants may have different effects on down-stream pathways, resulting in diverse phenotypic presentations. Furthermore, immunofluorescence analyses also indicated that certain variants led to protein mislocalization, underscoring the importance of proper nuclear localization for HDAC3’s function. It’s possible that the nuclear HDAC3 variants with compromised complex integrity may be more prone to ubiquitin degradation machinery, resulting in diminished levels of HDAC3 within the nucleus.

The six missense variants identified in this study fall into two distinct categories. The first group, including the D93N, P201S, L266S, and G267S variants, is located close to the enzyme’s active site, which may lead to decreased catalytic activities. We observed that individuals harboring the D93N and P201S variants, positioned slightly away from the active site compared to the L266S and G267S variants, exhibited relatively mild phenotypes. Accordingly, the individual 3, carrying the P201S variant, had only a single suspected seizure and abnormal EEG with generalized spike-wave and polyspike-wave complexes but not recurrent seizures or other features. In contrast, the second group, which includes the A110T and R359C variants, is not situated at the enzyme’s active site and thus retains intact catalytic activity when paired with the NCoR DAD domain. However, these variants resulted in decreased interactions with multi-protein complexes and altered nuclear localization. Particularly, individuals 1 and 2, carrying these variants, presented with pronounced phenotypes. Recently, Lacoste et al. reported that protein mislocalization, driven by effects on protein stability and membrane insertion, is the major mechanism for pathogenic missense variants.^35^ Collectively, our observations suggest that disrupted interactions with multi-protein complexes and the mislocalization of the HDAC3 complex may be a more critical mechanism of the observed phenotypes than the histone deacetylation activity itself.

The major clinical findings of HDAC3-related syndrome observed in our patients include neurodevelopmental disorder, congenital heart defects, and musculoskeletal abnormalities. Although previous studies have implicated these phenotypes in mice (MGI:1343091) using HDAC3 knock-out mouse models,^12–18^ our study further established a compelling link between *HDAC3* genetic variations and these phenotypes in humans. The other studies also reported additional phenotypes in mice, including disrupted lipid metabolism, hepatomegaly, and infertility.^36–38^ We did not observe these phenotypes in our patients—there was no clinical indication for specific assays to assess lipid metabolism and no evidence of increased liver size noted at the time when they presented for clinical care, and they are not at an age when infertility would have manifested. Future investigations within a larger cohort may provide further evidence on these phenotypic association in humans.

The *HDAC3* gene has a probability of loss-of-function intolerance (pLI) score of 1.00 and loss-of-function observed/expected upper bound fraction (LOEUF) score of 0.46 in the gnomAD v4.0.0 database,^39^ indicating that *HDAC3* is highly constrained and haploinsufficiency is likely the underlying mechanism. However, our experimental data suggest that not only diminished catalytic activity but also disrupted interactions with other multi-protein complexes and/or mislocalization of the complex constitute pivotal mechanisms. Therefore, it may be more complicated in *de novo* missense variants depending on the specific substitutions and the complexes they involve. To gain further insights into the underlying mechanisms and genotype-phenotype correlations, we analyzed cases with copy-number variations (CNVs) involving the *HDAC3* gene reported in the DECIPHER databases.^40^ This CNV analysis identified eleven patients with five large deletions and six large duplications, and most CNVs were suspected to have occurred *de novo* (**Figure S4**). Although phenotypic information was unavailable for three patients with large duplications, the other eight patients exhibited neurodevelopmental (7/8), skeletal (4/8), and cardiac (1/8) phenotypes, further supporting our findings.

The experimental findings underscore the pivotal role of the disrupted interaction with the CoREST complex, a shared mechanism among all HDAC3 variants. Notably, a Mendelian disorder linked to KDM1A (OMIM #609132)—an interacting partner of HDAC3 within the CoREST complex—has been recently described in three individuals.^20^ The clinical presentation in these individuals bore similarities to Kabuki syndrome, notably characterized by global developmental and speech delays, as well as distinctive facial features. The functional assessment of the identified KDM1A variants (E403K, D580G, and Y785H) revealed compromised catalytic activities and altered interactions with transcription factors.^41^ A phenotypic overlap was evident in our cohort with the KDM1A-associated disorder, suggesting a potential association with pathways related to the CoREST complex.

Another salient observation was the universally diminished interaction with AMER1 among all variants, as depicted in **Figure 3C**. Given that pathogenic variations in *AMER1* are established causes of Osteopathia striata with cranial sclerosis (OMIM #300373), manifesting in X-linked dominant inheritance, the diminished interaction with AMER1 might elucidate the musculoskeletal abnormalities observed in some patients. Similarly, diminished interactions were detected with *GANAB*, *AKAP8L*, and *EIF3B* across all examined variants (**Figure S5**). Prior reports have associated the gene dosage of *AKAP8L* with the microcephaly phenotype in humans^42,43^, suggesting that *AKAP8L* could play a role in the microcephaly phenotypes identified in our subjects. Future investigations focusing on these proteins are expected to offer a more comprehensive understanding of genotype-phenotype correlations for HDAC3-related syndromes.

Recent investigations offer valuable insights into potential therapeutic interventions for HDAC3-related syndrome. Watson et al.^26^ demonstrated that inositol phosphate analogues and derivatives, particularly those with Ins(1,4,5,6)P_4_, can activate the HDAC3:NCoR2 (SMRT) complex in the HDAC3 R265A variant. This activation has also been observed in the HDAC1 within the CoREST complex.^10^ Although these observations were not specifically made for HDAC3 in the CoREST complex, these findings imply that Ins(1,4,5,6)P_4_ could be a potential therapeutic candidate for HDAC3-related syndrome. It is imperative to conduct animal studies involving these agents, which might pave the way for innovative interventions in HDAC3-related syndrome.

In conclusion, this study presents evidence from six individuals with heterozygous *de novo* variants in *HDAC3*, accompanied by a spectrum of neurodevelopmental and other symptoms, supporting the role of *HDAC3* in human disease. These variants may disrupt interactions within multi-protein complexes, particularly those associated with the CoREST and NCoR complexes, and/or disrupt nuclear localization. Such disturbances could represent the primary pathogenic mechanisms underlying this syndrome. Further research involving a larger cohort of patients will be crucial to strengthen the connection between this gene and the disease, and to enhance our understanding of the genotype-phenotype relationships in HDAC3-related syndromes.

## Data Availability

The data supporting the findings of this study are available within the article and the Supplemental Material or can be made available upon reasonable request to the corresponding authors.

## Acknowledgments

This study was supported by the Institute of Information & Communications Technology Planning & Evaluation (IITP) grant, funded by the Korean government (MSIT, grant numbers: 2022-0-00333, RS-2023-00223069). Also, it was supported by the National Research Foundation of Korea (NRF) grants (NRF2021R1C1C1013220, NRF2022R1A5A102641311, and NRF2020R1A5A1019023), and the BK21 Four Biomedical Science Program. The SNUH Kun-hee Lee Child Cancer and Rare Disease Project Foundation, Republic of Korea (grant number: 22B-001-0100), the Research Resettlement Fund for the new faculty of Seoul National University, the Creative-Pioneering Researchers Program through Seoul National University, grants from Seoul National University College of Medicine, and the AI-Bio Research Grant through Seoul National University also supported this study.

## Author contributions

Conceptualization: J.G.Y., SK.L, JH.C, CH.L; Data Curation: S.L., J.C., HY.K., A.H.P., DDD study., SY.K., JM.K.; Investigation: J.G.Y., SK.L, H.S., D.H.; Supervision: JH.C, CH.L; Writing-original draft: J.G.Y., SK.L, CH.L; All authors reviewed and approved the final version of the manuscript.

## Supplemental information

Supplemental Figures 1–5, Supplemental Tables 1–2.

