## Supplemental information for "*De novo HDAC3* variants leading to epigenetic machinery dysfunction are associated with a neurodevelopmental disorder"

**< Table of Contents >**

**Supplementary Figures……………………………………………………………………..2**

**Fig. S1.** Confirmation of *de novo* HDAC3 variants using Sanger sequencing ....…………..……2

**Fig. S2.** Characterization of NCoR1 DAD domain-HDAC3 interaction and HDAC3 mutagenesis ……………………………………………………………………………………3

**Fig. S3.** Histone deacetylase activity of NCoR1 DAD domain-HDAC3 complex on acetylated mononucleosomes ..……..……………..............................................................................5

**Fig. S4.** Copy-number variations (CNVs) involving the *HDAC3* locus from the DECIPHER database ……………………………………………………………………………………..6

**Fig. S5.** Differential protein interaction profiles of HDAC3 variants in HEK293T cells ……......8

**Supplementary Tables…………………………………………………………….............9**

**Table S1.** Primer information ………………………………..……………………………..……9

**Table S2.** List of antibodies used in this study ……………………………..……..…………10

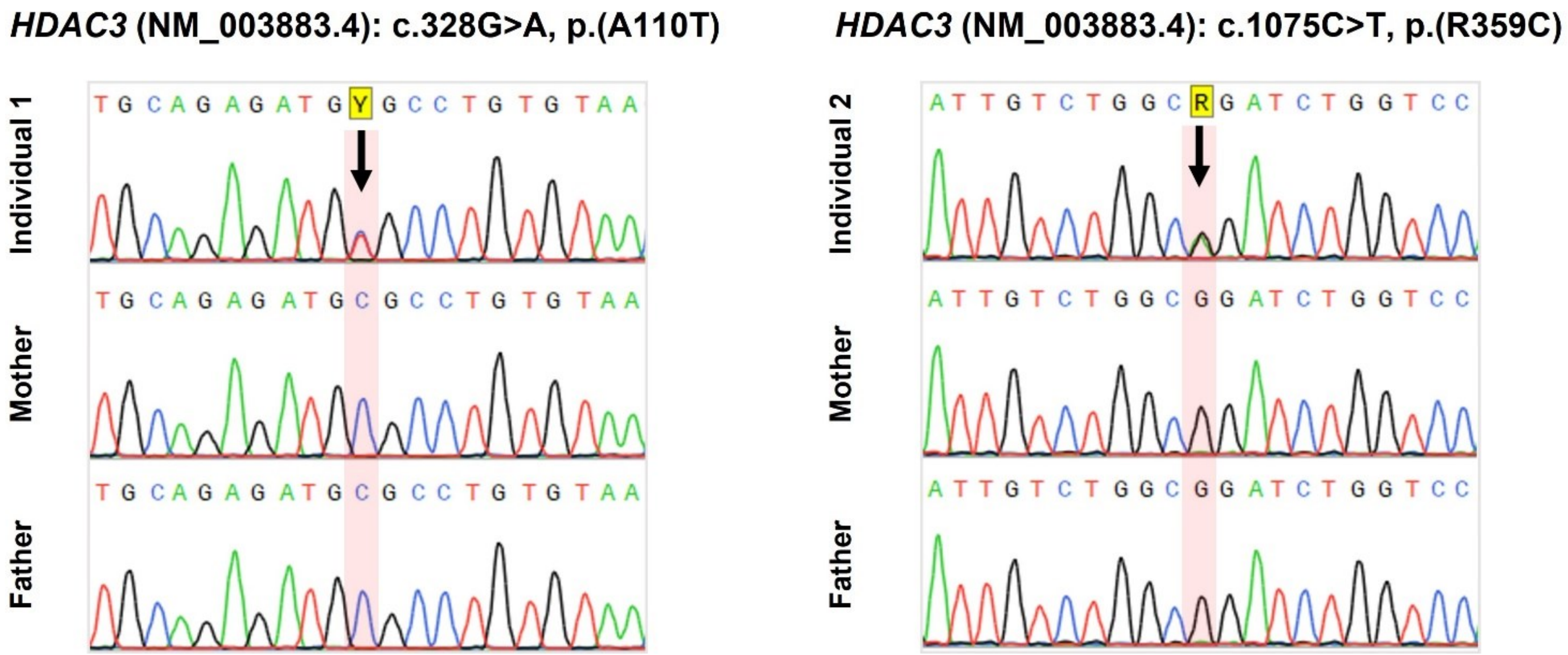


**Figure S1 | Confirmation of *de novo HDAC3* variants using DNA sanger sequencing**

Sanger sequencing was conducted to validate the *de novo* variants using the primers (**Table S1**) for individuals 1–2 and their parents. The results confirmed that the *HDAC3* variants occurred as *de novo*.

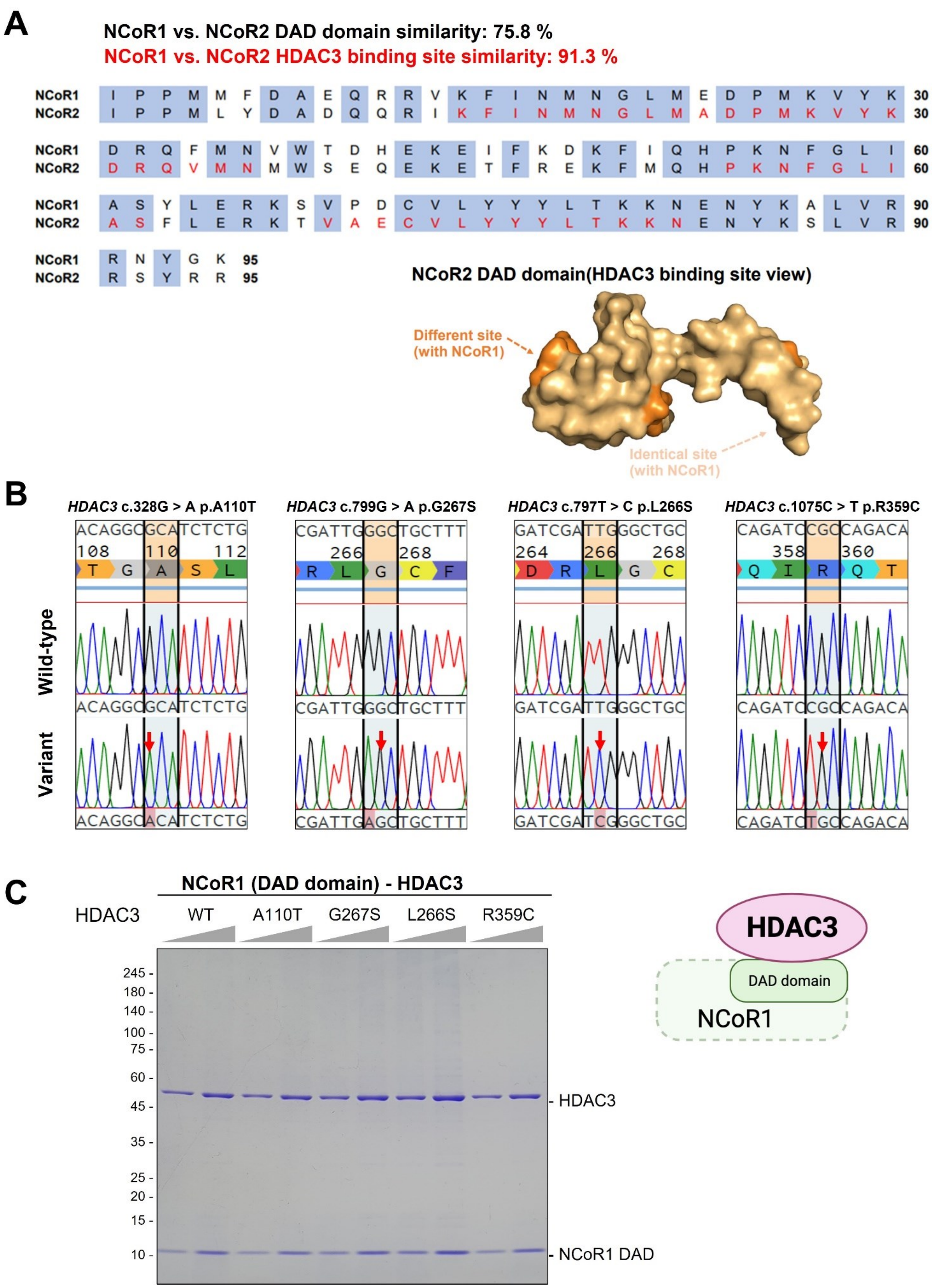
A
NCoR1 vs. NCoR2 DAD domain similarity: 75.8 %
NCoR1 vs. NCoR2 HDAC3 binding site similarity: 91.3 %
NCoR1 I P P M M F D A E Q R R V K F I N M N G L M E D P M K V Y K 30
NCoR2 I P P M L Y D A D Q Q R I K F I N M N G L M A D P M K V Y K 30
NCoR1 D R Q F M N V W T D H E K E I F K D K F I Q H P K N F G L I 60
NCoR2 D R Q V M N M W S E Q E K E T F R E K F M Q H P K N F G L I 60
NCoR1 A S Y L E R K S V P D C V L Y Y Y L T K K N E N Y K A L V R 90
NCoR2 A S F L E R K T V A E C V L Y Y Y L T K K N E N Y K S L V R 90
NCoR1 R N Y G K 95
NCoR2 R S Y R R 95
NCoR2 DAD domain(HDAC3 binding site view)
Different site (with NCoR1)
Identical site (with NCoR1)
B
HDAC3 c.328G > A p.A110T
HDAC3 c.799G > A p.G267S
HDAC3 c.797T > C p.L266S
HDAC3 c.1075C > T p.R359C
Wild-type
Variant
C
NCoR1 (DAD domain) - HDAC3
HDAC3
WT
A110T
G267S
L266S
R359C
245
180
140
100
75
60
45
35
25
20
15
10
HDAC3
NCoR1 DAD
HDAC3
DAD domain
NCoR1

### Figure S2 | Characterization of NCoR1 DAD domain-HDAC3 interaction and HDAC3 mutagenesis.

**A.** Sequence alignment of NCoR1 and NCoR2 DAD (Deacetylase Activating Domain) domains. This illustrates a 75.8% similarity in sequence, and a 91.3% similarity at the HDAC3 binding site. Alignment was conducted using the UniProt Align tool.

**B.** Mutagenesis of HDAC3. Four specific HDAC3 variants were introduced by PCR, utilizing mutagenic primers listed in Table S1. Sanger sequencing confirmed the presence of A110T, G267S, L266S, and R359C variants, indicating successful site-directed mutagenesis.

**C.** Purification of the NCoR1 DAD domain in complex with WT or HDAC3 variants. WT and variant forms of HDAC3, along with the NCoR1 DAD domain, were co-expressed in Sf9 cells using a baculovirus system. Complexes were purified using Ni-NTA Agarose and Anti-FLAG M2 Affinity Gel 64 hrs post-infection, as visualized on the SDS-PAGE gel, indicating no defects in their interaction with the NCoR1 DAD domain.

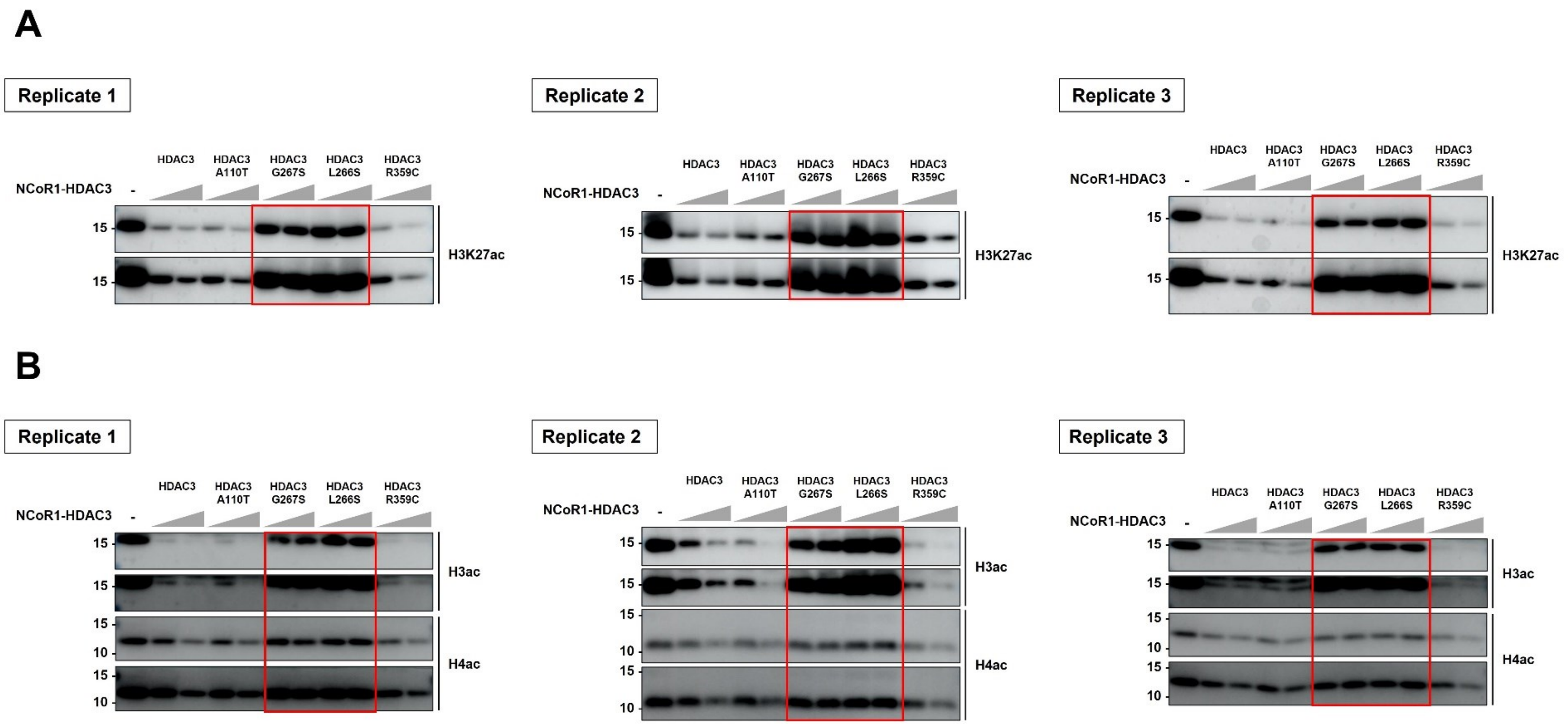


**Figure S3 | Histone deacetylase activity of NCoR1 DAD domain-HDAC3 complex on acetylated mononucleosomes.**

**A**. HDAC assay on H3K27ac mononucleosomes. This assay investigated the deacetylase activity of the NCoR1 DAD domain in complex with wild-type (WT) HDAC3 and its variants A110T, G267S, L266S, and R359C. Mononucleosomes acetylated at lysine 27 on histone H3 (H3K27ac) served as substrates. Immunoblotting with an H3K27ac-specific antibody quantified the deacetylation, revealing decreased activities in the G267S and L266S variants as denoted by the red boxes.

**B**. HDAC assay on H3ac/H4ac mononucleosomes. This panel assesses the deacetylase activities of the NCoR1 DAD domain-HDAC3 complex, both WT and variant forms, on mononucleosomes with acetylations at various lysine residues on histones H3 (H3K4,9,14,18ac) and H4 (H4K5,8,12,16ac). Antibodies specific to acetylated H3 (H3ac) and H4 (H4ac) detected the HDAC activities. The G267S and L266S variants exhibit notably reduced deacetylase activity, as highlighted in the red boxes across all replicates.

A

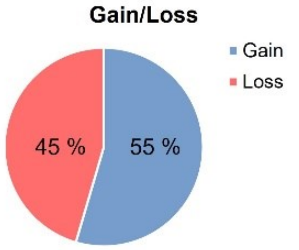


B

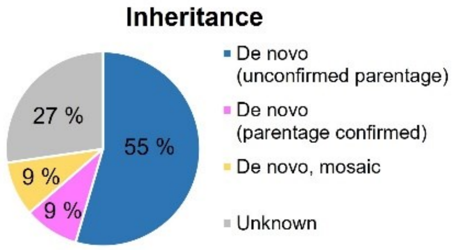


C

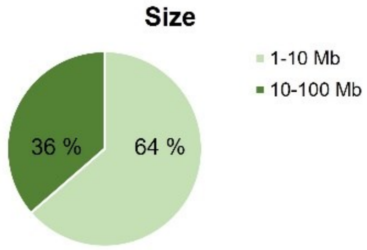


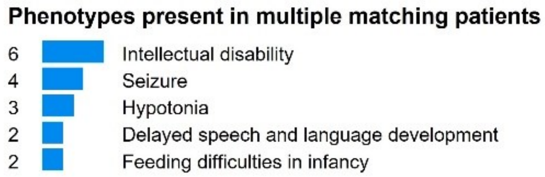


D

| Patient | Sex | Size | Type | Inheritance / Genotype | Phenotypes |
|---|---|---|---|---|---|
| **4681** | 46XX | 2.57 Mb | Deletion | Heterozygous de novo (unconfirmed parentage) | Delayed speech and language development; Feeding difficulties in infancy; Hypotonia; Intellectual disability; Seizure |
| **249028** | 46XX | 6.94 Mb | Deletion | Heterozygous de novo (unconfirmed parentage) | Abnormality of the upper respiratory tract; Coarse facial features; Hypotonia; Intellectual disability; Patent ductus arteriosus |
| **253734** | 46XY | 5.04 Mb | Deletion | Heterozygous de novo (unconfirmed parentage) | Feeding difficulties in infancy; Hypotonia; Intellectual disability |
| **255372** | 46XY | 17.93 Mb | Duplication | Heterozygous de novo (unconfirmed parentage) | 2-3 toe syndactyly; Inguinal hernia; Intellectual disability; Microcephaly; Sacral dimple; Seizure; Short stature |
| **261240** | 46XX | 24.76 Mb | Duplication | Heterozygous de novo (mosaic) | Intellectual disability; Seizure |
| **264075** | 46XX | 6.53 Mb | Deletion | Heterozygous de novo (unconfirmed parentage) | Abnormal plantar dermatoglyphics; Broad face; Lissencephaly; Seizure |
| **293879** | 46XX | 1.06 Mb | Duplication | Heterozygous (unknown) | NA |
| **307291** | 46XX | 2.96 Mb | Deletion | Heterozygous de novo (parentage confirmed) | Camptodactyly of finger; Eczema; Flexion contracture of toe; Hypohidrosis |
| **345239** | Unknown | 11.46 Mb | Duplication | Heterozygous (unknown) | NA |
| **401315** | 46XX | 9.06 Mb | Duplication | Heterozygous de novo (unconfirmed parentage) | Abnormal pinna morphology; Brachycephaly; Constipation; Deeply set eye; Delayed speech and language development; Diabetes mellitus; EEG abnormality; Fine hair; Gait disturbance; Hypertelorism; Intellectual disability; Mandibular prognathia; Precocious puberty in females; Prominent nose; Recurrent infections; Scoliosis; Short philtrum; Strabismus; Wide mouth |
| **473002** | 46XX | 15.94 Mb | Duplication | Heterozygous (unknown) | NA |

### Figure S4 | Copy-number variations (CNVs) involving the *HDAC3* locus from the DECIPHER database.

**A**. CNV gain and loss, **B**. inheritance pattern, **C**. size, and **D**. complete list of patients ($n$ = 11) involving the *HDAC3* locus reported in the DECIPHER database. Abbreviation: NA, not available.

**A**

**No. of altered interactions ($P$ < 0.05)**

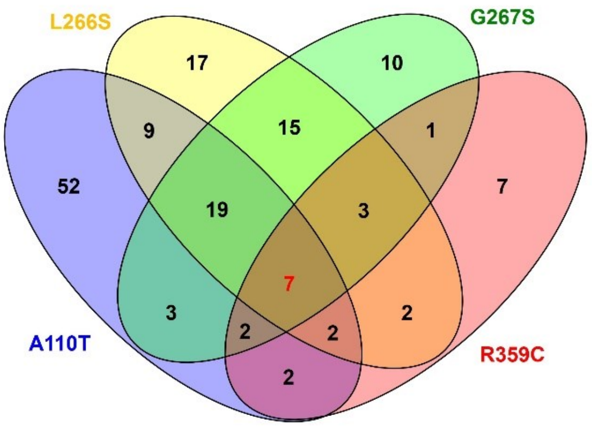


**B**

**Shared by all variants**

| Gene_name | Interaction |
|---|---|
| *KDM1A* | Down |
| *GANAB* | Down |
| *AMER1* | Down |
| *AKAP8L* | Down |
| *EIF3B* | Down |
| *HNRNPR* | Up |
| *HSPD1* | Down/Up |

**C**

**Shared by two or more variants: down-regulated GO term**

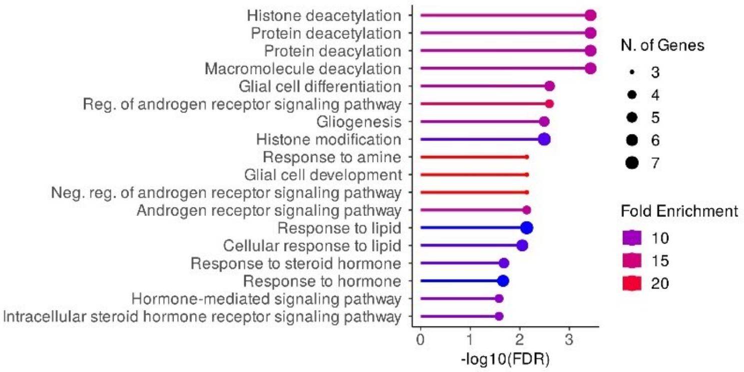


**D**

**Shared by two or more variants: up-regulated GO term**

| N | 10 |
|---|---|
| **GO category** | Proteasome complex (GO:0000502) |
| **Gene_name** | *HNRNPR, PSMA7, PSMB7, PSMC1, PSMC3, PSMC6, PSMD4, PSMD11, PSMD13, PSMD14* |

### Figure S5 | Differential protein interaction profiles of HDAC3 variants in HEK293T cells.

**A**. Venn diagram analysis. This panel presents a Venn diagram that quantifies the number of proteins exhibiting significantly altered interactions ($P$ < 0.05) with each of four *HDAC3* variants (A110T, L266S, G267S, and R359C) in comparison to the wild-type (WT) HDAC3 in HEK293T cells.

**B**. Consistent interaction changes across the *HDAC3* variants. This table enumerates specific proteins that display consistent changes in interaction levels across all *HDAC3* variants tested. The proteins *KDM1A*, *GANAB*, *AMER1*, *AKAP8L*, and *EIF3B* show uniformly reduced interactions across the variants, indicating a possible common pathway or functional disruption. In contrast, *HNRNPR*, which is associated with proteasome degradation pathways, exhibits an increased interaction.

**C**. Down-regulated GO terms. Bar chart of Gene Ontology (GO) terms enriched among proteins with decreased interaction with at least two variants, highlighting the involvement of histone deacetylation processes (GO:0016575), as expected.

**D**. Up-regulated GO terms. Most of up-regulated proteins, including the common involvement of *HNRNPR*, were participated in the proteasome complex (GO:0000502).

**Table S1 | Primer information.**

| Target | Primer | Sequences (5' to 3' end)[a] | Target size | $T_m$ |
|---|---|---|---|---|
| *HDAC3* (NM_003883.4): c.328G>A, p.(A110T) | Exon4_F | CCTAAGTCACAGTCCTTCCTGCC | 287 bp | 63.5 |
| | Exon4_R | ATGGAGATTGGAGGAATCTAGGATG | | 62.9 |
| *HDAC3* (NM_003883.4): c.1075C>T, p.(R359C) | Exon14_F | AGGTAAGCCAGAGGCAATTAAACT | 383 bp | 60.8 |
| | Exon14_R | CTGAACTAGAGGTACCACTGAGATG | | 58.6 |
| A110T mutagenesis | A110T_F | CTCTTTGAGTTCTGCTCGCGTTACACAGGCACATCTCTGCAAGGAGCAACCCAGCTGAACAAC | - | 76.8 |
| | A110T_R | GTTGTTCAGCTGGGTTGCTCCTTGCAGAGATGTGCCTGTGTAACGCGAGCAGAACTCAAAGAG | | 76.8 |
| G267S mutagenesis | G267S_F | TGTGGAGCTGACTCTCTGGGCTGTGATCGATTGAGCTGCTTTAACCTCAGCATCCGAGGGCATGGG | - | 80.4 |
| | G267S_R | CCCATGCCCTCGGATGCTGAGGTTAAAGCAGCTCAATCGATCACAGCCCAGAGAGTCAGCTCCACA | | 80.4 |
| L266S mutagenesis | L266S_F | TGTGGAGCTGACTCTCTGGGCTGTGATCGATCGGGCTGCTTTAACCTCAGCATCCGAGGGCATGGG | - | 82.3 |
| | L266S_R | CCCATGCCCTCGGATGCTGAGGTTAAAGCAGCCCGATCGATCACAGCCCAGAGAGTCAGCTCCACA | | 82.3 |
| R359C mutagenesis | R359C_F | CAGAACTCACGCCAGTATCTGGACCAGATCTGCCAGACAATCTTTGAAAACCTGAAGATGCTG | - | 74.8 |
| | R359C_R | CAGCATCTTCAGGTTTTCAAAGATTGTCTGGCAGATCTGGTCCAGATACTGGCGTGAGTTCTG | | 74.8 |

[a]Mutated nucleotide sites are highlighted in red for mutagenic primers.

**Table S2 | List of antibodies used in this study.**

| Category | Name | Source | Cat No. |
|---|---|---|---|
| **Primary antibody** | | | |
| | FLAG M2 | Sigma-Aldrich | F1804 |
| | GAPDH | GeneTex | GTX627408 |
| | HDAC3 | Cell Signaling Technology | 85057 |
| | NCoR1 | Cell Signaling Technology | 5948 |
| | NCoR2/SMRT | Cell Signaling Technology | 62370 |
| | KDM1A/LSD1 | Cell Signaling Technology | 2184 |
| | β-tubulin | Abbkine | A01030 |
| | H3K9ac | Cell Signaling Technology | 9649 |
| | H3K27ac | Abcam | ab4729 |
| | H3ac | Active Motif | 39139 |
| | H4ac | Active Motif | 39243 |
| | H3 | Abcam | ab1791 |
| | H4 | Abcam | ab10158 |
| **Secondary antibody** | | | |
| | Goat anti-mouse IgG F(ab'), polyclonal antibody (HRP conjugate) | Enzo Life Science | ADI-SAB-100-J |
| | Goat anti-rabbit IgG, polyclonal antibody (HRP conjugate) | Enzo Life Science | ADI-SAB-300-J |
| | Anti-mouse IgG-Alexa Fluor 488 secondary antibody | Invitrogen | A-11001 |
